# Transparent comparisons of Emergency-Department prioritization policies: integrating tail risk, target attainment, and utility analysis

**DOI:** 10.1101/2025.06.05.25329094

**Authors:** Adam DeHollander, Mark Karwan, Sabrina Casucci

## Abstract

Comparative studies of emergency-department (ED) patient-prioritization rules frequently rely on single, average-based metrics, obscuring clinically important trade-offs. We introduce a three-part evaluation framework that combines tail-sensitive summary statistics, threshold-attainment curves, and stakeholder-informed utility analysis, and apply it in a discrete-event simulation of a 30-bed mixed-acuity ED. Tail statistics expose extremes—for example, the 99th-percentile length-of-stay (LOS) gap between strategies exceeds ten times the corresponding mean gap—and reveal cohort-specific inequities masked by overall averages. Threshold-attainment curves map the fraction of patients meeting every LOS target, showing where strategy rankings switch as service standards tighten or relax. Utility contours translate multi-cohort performance into a single stakeholder score, demonstrating how preferred rules pivot when institutional priorities shift from rapid discharge of low-acuity patients to protection of higher-acuity throughput. Together, the techniques uncover complementary, non-redundant information: each one highlights strengths and weaknesses invisible to the others. The framework therefore offers a transparent, replicable template for selecting prioritization policies that align with local clinical objectives, resource constraints, and risk tolerances.

## 1 Introduction and Literature Review

### 1.1 Background and Significance

Emergency-department (ED) crowding remains a persistent threat to timely, high-quality care, with over 90% of EDs reporting regular crowding as of 2016—a situation further exacerbated by the COVID-19 pandemic [1]. Prolonged length of stay (LOS), boarding, and repeated “left-without-being-seen” (LWBS) events have each been linked to excess mortality, lower patient satisfaction, and staff burnout [2, 3]. Long wait times not only delay time-sensitive interventions for high-acuity patients but also deteriorate the overall quality of care [4–7]. Even low-acuity patients may experience prolonged discomfort or clinical deterioration if neglected. Beyond clinical outcomes, crowding constrains a hospital’s ability to respond to new emergencies and impairs operational efficiency, with downstream effects on financial performance [8]. While root causes span hospital-wide capacity constraints, one operational lever squarely under the control of ED managers is the patient-prioritization strategy—that is, the rule that determines which patient is served next once a resource becomes available. Over the past two decades, researchers have proposed a rich catalog of such rules, ranging from simple first-come-first-served (FCFS) queues to dynamic algorithms that blend acuity, projected workload, and downstream bed availability [9].

Despite this methodological progress, evidence on *which* strategy works best is inconclusive. Primary studies differ widely in (i) the key-performance indicators (KPIs) they report (e.g., mean LOS versus 90th-percentile LOS), (ii) whether they examine distribution tails, and (iii) the degree to which stakeholder preferences are incorporated. Inconsistent evaluation protocols lead to the paradox that the *same* strategy can appear superior under one article’s metrics yet inferior under another’s, clouding generalizability and hindering adoption [10]. The absence of a systematic evaluation framework therefore represents both a scientific gap and a practical barrier to evidence-based ED operations.

This paper addresses that gap by introducing a unified framework built on three complementary evaluation techniques. The first technique involves computing KPI summary statistics with explicit tail analysis to detect hidden extremes in performance distributions. The second constructs threshold-based performance curves that reveal sensitivity to time-target selection, making it easier to interpret operational trade-offs. The third incorporates stakeholder-informed utility functions that translate multidimensional clinical objectives into a single, interpretable scalar score.

Using nine prioritization rules—including widely studied approaches (e.g., FCFS, Accumulating Priority Queue [APQ]) and several novel strategies—we demonstrate that each rule reveals distinct trade-offs that are obscured by mean-only performance metrics. We show that a strategy may appear optimal under one evaluation criterion but perform poorly under another. Our aim is not to identify a single “best” rule, but to equip ED decision-makers with a transparent framework for comparing strategies within their specific operational contexts.

### 1.2 Literature Review

#### 1.2.1 Patient-flow challenges in crowded EDs

Systematic reviews consistently find that crowding worsens clinical outcomes and elevates operational costs [3, 4, 8]. Among the most frequently reported indicators are median length of stay (LOS) and time-to-physician [3, 10]. However, these studies also emphasize that extreme tail events—such as 99th-percentile LOS—are more often responsible for serious incidents like ambulance diversions than average performance metrics.

#### 1.2.2 Patient-prioritization strategies

A wide array of interventions have been proposed to alleviate emergency department (ED) crowding, including the use of telehealth solutions [4], educational initiatives for patients, and process-improvement frameworks like Six Sigma [11]. This review, however, centers on patient prioritization strategies—rules that dictate the sequencing of patients when a treatment resource becomes available. The most basic of these is the first-come, first-served approach, while more advanced methods utilize algorithmic or heuristic logic to improve performance metrics. Prior work has explored a diverse spectrum of prioritization methods, spanning from static mechanisms such as structured priority queues [12–14] to adaptive systems that make real-time decisions [15–17]. Some authors have pursued optimization-based formulations [18, 19], although these often struggle with the incorporation of uncertainty. Other researchers have turned to data-driven techniques like machine learning to better model and respond to stochastic dynamics [20, 21]. Additional investigations have examined more traditional strategies, including revised triage procedures [9, 22] and evaluations of heuristic decision-making by frontline clinicians [23, 24]. Comparative studies typically employ discrete-event simulation because it provides patient-level counterfactuals without disrupting care [25].

#### 1.2.3 Existing evaluation practices

Although numerous studies investigate patient prioritization strategies, no standard approach exists for evaluating their effectiveness. Instead, three methodological traditions have emerged in the literature.

The first and most common approach involves reporting single-moment KPIs, such as mean or median length of stay (LOS), wait time, or throughput [25–28]. This practice is widespread in both simulation and empirical studies. However, since LOS distributions are typically right-skewed, focusing on central moments alone can obscure rare but operationally severe delays.

A second approach assesses target-achievement rates, often in alignment with regulatory benchmarks— such as the proportion of patients discharged within a 4-hour window [29]. While such metrics are straightforward to interpret and align with policy goals, they depend heavily on the chosen threshold, which may be arbitrary and insensitive to broader performance variation.

The third approach, found in a smaller body of literature, employs utility-based multicriteria scoring to synthesize performance across several KPIs. These studies use explicit utility functions—linear, exponential, or Chebyshev—to represent stakeholder preferences [16, 30]. Although this method enhances transparency, most implementations do not test the robustness of results to changes in utility-parameter values, limiting their prescriptive reliability.

#### 1.2.4 Lack of generalizability

Early evidence suggested that simply introducing a structured triage scale would shorten waits and lower mortality [31]. Yet subsequent observational work uncovered the opposite effect: Sax *et al*. noted that widespread assignment to ESI Level 3 created a “mid-acuity log-jam,” lengthening throughput for all but the sickest patients because beds were occupied by patients whose severity had been overestimated [32]. These conflicting findings highlight that the same triage rule can either alleviate or exacerbate crowding.

Commentaries have questioned whether current validation methods transfer across jurisdictions. Twomey *et al.* argued that techniques developed in well-resourced settings “may not be appropriate and repeatable in developing countries,” and highlighted conceptual problems in declaring any single metric the gold standard for validity [33]. One hospital might benchmark triage accuracy against ICU admission; another might use expert consensus. Such heterogeneity makes cross-site comparison—and thus generalizability— elusive.

The same pattern emerges in the rapidly growing AI/ML triage literature. El Arab and Al Moosa found that most machine-learning studies were single-center and lacked external validation, with selection bias and overfitting as recurrent threats [34]. When Ryu *et al.* trained a gradient-boosted triage score at one hospital and deployed it at sister sites, the AUC for predicting admission ranged from 0.93 to 0.71 across locations served by the same health system [35]. Broader reviews of data-driven admission predictors echo the call for “rigorous external evaluation before clinical use” [36]. Meanwhile, Ingielewicz *et al.* surveyed traditional scales and concluded that “no existing triage system clearly outperforms others in every aspect,” effectively dispelling the notion of a universal best-in-class tool [37]. From a resource-constrained perspective, Siddiqui *et al.* stated that “the need and practical applicability of any triage is dictated by the hospital system and setting” [38].

Compounding these issues, most primary studies compare a candidate rule only to baseline practice rather than to other advanced rules, and they often rely on a single evaluation metric [20, 22]. As we later demonstrate, the ranking of nine common prioritization strategies changes when analysts shift from mean LOS to tail-sensitive or utility-based criteria within the same 30-bed ED. If evaluation choice alone can flip conclusions in one configuration, then extrapolating results across hospitals with different capacity, acuity mix, or stakeholder priorities is doubly precarious.

#### 1.2.5 Gaps in the literature

To our knowledge, **no consensus exists** on which KPIs constitute the minimal reporting set when analyzing a patient prioritization strategy. Recent umbrella reviews explicitly call for “standardized, multidimensional evaluation frameworks” to enable meta-analysis and real-world translation [10].

### 1.3 Contributions and Organization

Responding to these calls, we synthesize and extend prior work by formalizing three mutually reinforcing techniques that together (i) oblige analysts to quantify tail behavior, (ii) expose threshold dependence, and (iii) embed stakeholder utilities without collapsing them into opaque composite indices. By demonstrating the framework on nine prioritization rules within a unified simulation environment, we provide the first head-to-head illustration of how evaluation choice alone can reverse strategic rankings—thereby underscoring the necessity of consistent, transparent methodologies.

The remainder of the study is organized as follows. Section 2 details the proposed methodology, including cohort-specific metrics, threshold-attainment curves, and utility analyses; Section 3 presents illustrative results from a calibrated 30-bed ED simulation; Section 4 discusses managerial implications, limitations, and avenues for future research; and Section 5 concludes by summarizing the key contributions, offering practical recommendations for aligning prioritization rules with clinical objectives, and highlighting the study’s broader significance. Supplementary details appear in the Appendices: Appendix A describes each prioritization strategy; Appendix B defines the stakeholder utility functions; Appendix C provides full simulation-model parameters; and Appendix D presents the context-specific formulation of the area under the curve (AUC) that is used in our threshold-attainment analysis in Section 2.3.

## 2 Materials and Methods

### 2.1 Study design and objectives

We propose an evaluation framework that compares ED patient-prioritization strategies along three complementary dimensions: distributional tail risk, threshold attainment, and stakeholder-aligned utility. This framework addresses the inconsistencies outlined in Section 1.2.4. All strategies are evaluated using a common discrete-event simulation (DES) model calibrated to a 30-bed, mixed-acuity ED. Implementation details, utility-function specifications, and full simulation parameters are provided in Appendices A–D to avoid diverting focus from the primary contribution of this work and to improve the flow of the paper to benefit the reader. Although we introduce novel strategies and report illustrative results, these are intended solely to demonstrate the application of the evaluation methods. The core aim of this study is to advance the methodology for comparing patient prioritization strategies, not to advocate for specific strategies.

Emergency departments monitor a range of key performance indicators—door-to-doctor time (DTDT), the proportion who leave without being seen (LWBS), mortality rates, patient-reported satisfaction, and others—each reflecting different aspects of crowding [3]. We restrict our analysis to length of stay (LOS) because it is both the most reported metric and one with clear relevance to clinicians, administrators, and patients. The proposed evaluation framework is agnostic to the chosen KPI and can be replicated with any of these measures; expanding the study to multiple indicators would chiefly lengthen the manuscript without enriching its methodological contribution as our intent is to showcase the framework, not to advocate a specific prioritization rule. Moreover, High-acuity (ESI 1) arrivals are excluded from all subsequent analyses because, regardless of patient prioritization strategy, these patients receive immediate treatment.

Finally, Table 1 summarizes the notation we use throughout this manuscript.

**Table 1.** Notation primer. Table 1 summarizes symbols that re-occur throughout Sections 2–3.

| Symbol | Meaning |
| --- | --- |
| $P$ | cohort under study (e.g., all patients, low-acuity) |
| $M_i$ | KPI value for patient $i \in P$ |
| $T_P(t)$ | proportion of cohort $P$ with $M_i \leq t$ |
| $t^{max}$ | maximum threshold considered |
| $t_X$ | time at which $T_P(t) = X/100$ |
| $U(\cdot)$ | stakeholder-informed utility score |

### 2.2 Technique 1: KPI Summary Statistics and Tail Analysis

This technique reflects standard practice by summarizing the distribution of each KPI. It is intentionally simple and does not incorporate advanced methodological tools. We include it because many studies provide limited distributional insight, typically reporting only the mean and occasionally the standard deviation. In contrast, our approach reports both central-tendency and right-tail metrics, as the extreme upper tail of the KPI distribution often corresponds to the most critical and undesirable outcomes—those that pose the greatest risks to patients and operational performance—but is seldom addressed in the literature.

Assuming that lower KPI values indicate better performance, we recommend reporting a comprehensive set of statistics for each strategy. These include the sample size, which ensures transparency about the number of observations and supports the assessment of statistical power; the mean, which reflects the average outcome and facilitates comparison of overall performance; and the median, which serves as a robust measure of central tendency that is not overly influenced by outliers. Additionally, reporting the minimum and maximum values allows for identification of the best and worst observed outcomes, respectively—offering insight into exceptional performance as well as potential failures.

To quantify tail risk explicitly, we also recommend including the 75th, 90th, 95th, and 99th percentiles of each KPI distribution. These percentiles reveal how frequently patients experience extremely long waits or lengths of stay, offering a granular view of performance in high-risk scenarios. This practice is consistent with prior guidelines in emergency department analytics [3].

Each of these metrics can—and should—be reported separately for relevant cohorts. In Section 3 (Results), for example, we will present these statistics for all patients combined, as well as stratified by Low and Mid acuity groups, which will reveal differing performance profiles when disaggregated. At a minimum, reports should be stratified by acuity level; additional cohort definitions might include arrival-time window, required resource type, boarding status, or other clinically meaningful categories.

### 2.3 Technique 2: Threshold-Based Performance

While summary statistics and tail percentiles (Technique 1) reveal overall distributional properties, stakeholders often specify explicit time targets—for example, the proportion of patients discharged within four hours. Technique 2 addresses this by quantifying, for each strategy and cohort, the fraction of patients whose KPI falls below a clinically meaningful threshold. By evaluating this performance over a range of thresholds, analysts can visualize and compare how sensitive each prioritization rule is to the choice of time target.

We begin by defining the indicator function

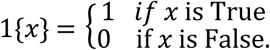

Let *P* denote a patient cohort (e.g., all patients or those in a given acuity group), and let *M*_*i*_be the KPI value for patient *i* ∈ *P* (e.g., length of stay, waiting time, door-to-doctor time, boarding indicator, or satisfaction score). For a specified threshold *t*, we define the threshold-attainment function

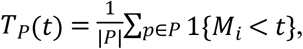

which represents the proportion of patients in cohort *P* whose KPI does not exceed *t*.

In practice, we compute *T*_*P*_(*t*) over a grid of thresholds *t* ∈ {*t*_1_, *t*_2_,⋯,*t*_*K*_ }. Plotting *T*_*P*_(*t*) against *t* yields a threshold-attainment curve, with higher curves indicating faster achievement of the target. Separate curves are generated for each strategy *s* and cohort *P*, enabling direct visual comparison. Confidence bands (e.g., bootstrap 95% intervals) may be overlaid to assess statistical significance.

For our illustrative analysis, we compute *T*_LOS―Low_(*t*) and *T*_LOS―Mid_(*t*) where *P* corresponds to the low- and mid-acuity groups, respectively, and *M*_*i*_ is each patient’s length of stay. By comparing these curves across the nine prioritization rules, one can readily identify how many patients meet a four-hour discharge target and observe how performance changes under more stringent (e.g., three-hour) or more lenient (e.g., five-hour) thresholds.

Threshold-attainment analysis offers several key advantages. First, plotting curves over a continuous range of *t* facilitates a sensitivity analysis, since intersections of curves reveal threshold ranges in which one strategy outperforms another. Second, curves directly encode clinical relevance, allowing stakeholders to read off the proportion of patients meeting an institution’s policy-driven time target without recomputing separate summary statistics. Third, when it is impractical to display full curves—such as in print-constrained venues—selecting a few representative percentile targets (for instance, the time to 75 percent, 90 percent, or 95 percent attainment) and tabulating those values can achieve space efficiency.

Beyond visual inspection, several summary metrics can be derived from *T*_*P*_(*t*) to facilitate single-value comparisons. The **area under the curve (AUC)** over the interval [0, *t*^max^] is defined by

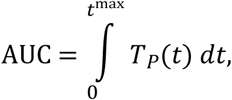

or more generally

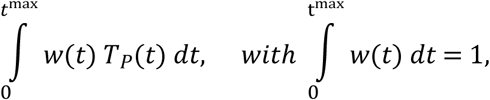

where *w*(*t*) is a weight function (e.g., linear or exponential decay) emphasizing early, middle, or late thresholds and *t*^*max*^represents the upper bound of interest (such as 12 hours for LOS). To standardize comparisons across KPIs, we report the standardized AUC as *AUC*/*t*^*max*^, which ranges from 0 to 1. Appendix D provides the full details of our AUC calculations. AUC is particularly beneficial because it consolidates performance across the entire range of clinically relevant thresholds into a single measure, enabling straightforward quantitative comparison of different strategies across all thresholds.

A second concise metric is the **speed to** ***X***% **attainment**, defined by

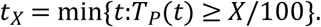

which answers the question, “How long until *X*% of patients meet the KPI target?” For example, one might report that “Strategy A reaches 90 percent discharge 25 minutes faster than Strategy B for mid-acuity patients.” Because *X* is a stakeholder-defined parameter, it can be varied to reflect different operational goals.

Researchers can tabulate AUC or *t*_*X*_values for key percentiles—such as 50 percent, 75 percent, 90 percent, and 95 percent—to present succinct comparisons without plotting every curve. All threshold-based metrics—full curves, AUC, and *t*_*X*_—should be reported separately for each cohort of interest. In Section 3 (Results), we will present these metrics for low-acuity and mid-acuity groups, demonstrating how strategy rankings may shift when performance is disaggregated. At a minimum, stratification by acuity level is required; additional analyses may consider other cohorts (for example, arrival-time windows, required resource types, or boarding status) to uncover subgroup-specific trade-offs.

### 2.4 Stakeholder-Informed Utility Functions

Techniques 1 and 2 quantify *what* each prioritization rule achieves in terms of KPI distributions and threshold attainment, but they do not capture *how much* value stakeholders assign to those outcomes. Technique 3 remedies this by mapping multi-dimensional performance into a single, stakeholder-aligned score, *U*( · ), constructed to increase as outcomes improve. While the precise functional form—whether linear, quadratic, exponential, Chebyshev, or otherwise—is specified in Appendix B, the following paragraphs describe how utility-based visualizations enhance decision support and how we apply them in our illustrative Results. The numerical utility parameters used in our analysis are illustrative placeholders; they were not elicited from ED stakeholders and serve only to demonstrate Technique 3.

First, one can plot

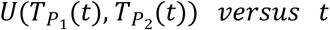

for each strategy where *P*_1_ and *P*_2_ are two cohorts. In our illustrative analysis, we use *U*(*T*_LOS―Low_(*t*),

*T*_LOS―Mid_(*t*)). By tracking utility across the same grid of LOS thresholds used in Technique 2, these curves reveal trade-offs in a single view: a rule that underperforms at short thresholds may nonetheless deliver high stakeholder value at longer thresholds, indicating favorable tail performance despite mediocre central metrics.

Second, for any fixed threshold *t*^∗^, a threshold-specific utility scatter plot provides an intuitive and easy to interpret visualization. We plot

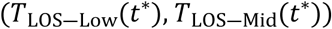

on the horizontal and vertical axes, respectively, and overlay utility contours *U*( · ) = *c*. Each strategy appears as a point colored by its utility level (e.g., green for high, red for low). Note that both axes remain raw KPI percentages; utility influences only the contour lines and point colors. This visualization combines the interpretability of familiar KPI percentages with a direct encoding of stakeholder preferences.

Finally, when multiple utility formulations or hyperparameter settings are under consideration, a cross-utility comparison can be performed by plotting *U*_1_against *U*_2_for each strategy at selected thresholds. Near-linear alignment along a positively sloped line indicates that strategic ordering is robust to the choice of utility specification; divergence suggests that different stakeholder weighting induces materially different recommendations, warranting closer examination.

These utility-driven plots enrich our evaluation framework by embedding explicit stakeholder trade-offs into familiar performance metrics, highlighting threshold dependence—since the utility curves mirror LOS threshold curves while summarizing value rather than raw attainment—and testing robustness through cross-utility comparisons that guard against overconfidence in any single preference model.

## 3 Results

All results in this section are **illustrative**—they reflect a single discrete-event-simulation calibration for a 30-bed mixed-acuity ED and utility parameters chosen for demonstration only. They should not be interpreted as prescriptive guidance for any specific hospital.

### 3.1 KPI summary statistics and tail analysis

Table 2 presents LOS summary statistics for the combined low- and mid-acuity cohort. Across the nine prioritization strategies (detailed in Appendix A), central tendency differs minimally—the grand mean ranges only seven minutes (193 to 200 min)—but the right-hand tail varies substantially, with a 99th-percentile spread of approximately 59 minutes.

**Table 2.** Length-of-Stay Summary Statistics by Prioritization Strategy and Acuity Cohort. This table presents key descriptive and tail-focused metrics for patient length of stay (LOS) under nine prioritization rules in a simulated 30-bed mixed-acuity emergency department. Columns report the number of observations, mean, median, 75th, 90th, 95th, and 99th percentiles, standard deviation, and observed minimum and maximum. Results are shown first for the combined low- and mid-acuity cohort (“Overall”), then separately for low-acuity and mid-acuity patients. The narrow range of grand means (193–200 min) contrasts sharply with the 99th-percentile spreads—59 min overall, 102 min for low acuity, and 116 min for mid acuity—highlighting how tail behavior diverges across strategies and cohorts.

| Strategy | Count | Mean | Median | P75 | P90 | P95 | P99 | STD | Min | Max |
| --- | --- | --- | --- | --- | --- | --- | --- | --- | --- | --- |
| **Overall** |  |  |  |  |  |  |  |  |  |  |
| AAPQ-LWP-PFT | 12377 | 195 | 191 | 234 | 280 | 326 | 459 | 81 | 32 | 968 |
| AAPQ-LWP | 12602 | 195 | 193 | 231 | 277 | 325 | 481 | 82 | 32 | 956 |
| AAPQ-PFT | 12671 | 197 | 189 | 237 | 293 | 349 | 473 | 86 | 32 | 1050 |
| AAPQ | 12484 | 195 | 190 | 235 | 287 | 335 | 450 | 81 | 32 | 1127 |
| Acuity-Based FCFS | 12578 | 200 | 203 | 260 | 308 | 350 | 469 | 95 | 32 | 845 |
| APQ | 12649 | 197 | 186 | 233 | 295 | 351 | 484 | 86 | 32 | 1185 |
| FCFS | 12108 | 195 | 181 | 229 | 305 | 351 | 425 | 79 | 32 | 1148 |
| LWP | 12502 | 193 | 187 | 227 | 269 | 306 | 442 | 76 | 32 | 1247 |
| PFT | 12008 | 198 | 193 | 239 | 286 | 329 | 442 | 78 | 32 | 1531 |
| **Low Acuity** |  |  |  |  |  |  |  |  |  |  |
| AAPQ-LWP-PFT | 9266 | 192 | 190 | 230 | 270 | 301 | 374 | 70 | 32 | 948 |
| AAPQ-LWP | 9443 | 192 | 192 | 228 | 266 | 301 | 404 | 72 | 32 | 956 |
| AAPQ-PFT | 9501 | 188 | 184 | 226 | 268 | 310 | 407 | 74 | 32 | 737 |
| AAPQ | 9381 | 188 | 187 | 227 | 270 | 306 | 393 | 71 | 32 | 831 |
| Acuity-Based FCFS | 9424 | 190 | 198 | 253 | 294 | 320 | 385 | 87 | 32 | 707 |
| APQ | 9489 | 184 | 179 | 216 | 262 | 310 | 427 | 75 | 32 | 1185 |
| FCFS | 9134 | 170 | 168 | 197 | 232 | 258 | 325 | 55 | 32 | 834 |
| LWP | 9372 | 187 | 185 | 220 | 259 | 287 | 370 | 66 | 32 | 1105 |
| PFT | 9003 | 190 | 187 | 230 | 270 | 305 | 387 | 71 | 32 | 732 |
| **Mid-Acuity** |  |  |  |  |  |  |  |  |  |  |
| AAPQ-LWP-PFT | 2668 | 189 | 173 | 232 | 301 | 371 | 534 | 100 | 42 | 930 |
| AAPQ-LWP | 2694 | 196 | 176 | 238 | 321 | 397 | 599 | 108 | 41 | 882 |
| AAPQ-PFT | 2710 | 213 | 199 | 267 | 349 | 411 | 546 | 109 | 40 | 1050 |
| AAPQ | 2648 | 203 | 191 | 254 | 331 | 392 | 497 | 101 | 37 | 1127 |
| Acuity-Based FCFS | 2693 | 218 | 206 | 272 | 358 | 419 | 573 | 109 | 42 | 845 |
| APQ | 2694 | 227 | 216 | 278 | 355 | 407 | 554 | 102 | 45 | 911 |
| FCFS | 2523 | 263 | 257 | 318 | 367 | 395 | 483 | 87 | 45 | 1148 |
| LWP | 2668 | 205 | 187 | 250 | 305 | 364 | 538 | 99 | 45 | 1247 |
| PFT | 2565 | 214 | 201 | 264 | 328 | 375 | 492 | 94 | 43 | 1531 |

When stratified by acuity level, sharper contrasts emerge. For low-acuity patients, the mean LOS range triples to 22 minutes (170 to 192 min) and the 99th-percentile spread widens to 102 minutes. In contrast, mid-acuity patients exhibit a mean range of 74 minutes (189 to 263 min) and a 99th-percentile spread of 116 minutes.

Acuity stratification also reverses strategy rankings. For low-acuity patients, FCFS outperforms all other rules on mean, median, and percentile metrics, whereas AAPQ-LWP-PFT yields the highest mean LOS. In the mid-acuity cohort, FCFS performs worst in mean, median, and upper-tail percentiles, while AAPQ-LWP-PFT ranks best on those metrics.

However, rankings vary by KPI. Among low-acuity patients, AAPQ-LWP-PFT produces the worst mean (albeit by a narrow margin) yet its right-tail performance remains near average. For mid-acuity patients, AAPQ-LWP excels on mean, median, and 75th-percentile metrics but performs poorly in more extreme tail measures, such as the 95th and 99th percentiles.

These findings underscore two key points: averaging across acuity levels conceals clinically meaningful variation, and tail-focused metrics often contradict conclusions based on central tendency alone.

### 3.2 Threshold-based performance

Figure 1 plots the percentage of low-acuity patients discharged within *t* minutes, denoted *T*_LOS―Low_(*t*). Several patterns emerge. For short thresholds (*t* < 120 minutes), Acuity-Based FCFS outperforms other strategies by approximately 10 percentage points; however, its relative performance deteriorates at higher thresholds (*t* > 200 minutes). Beyond this, the choice of threshold does not meaningfully affect the relative ordering of most strategies.

**Figure 1.** Cumulative Discharge Profiles for Low-Acuity Patients. This figure plots T̂_LOS―Low_(t), the percentage of low-acuity patients discharged within t minutes, for each prioritization strategy over a 12-hour window. The early-time advantage of Acuity-Based FCFS is evident for t < 120 min—outperforming alternatives by roughly 10 percentage points—while its performance converges or declines relative to other rules at longer thresholds.

Analogous trends appear for mid-acuity patients in Figure 2, where the curves similarly suggest minimal divergence between strategies across thresholds. These results imply that the choice of threshold, in such contexts, is unlikely to affect decision-making. Notably, Figure 2 also reinforces the pattern observed in Section 3.1: FCFS underperforms during the initial 5 hours but surpasses other strategies at later thresholds.

**Figure 2.** Cumulative Discharge Profiles for Mid-Acuity Patients. This figure presents T̂_LOS―Mid_(t), the share of mid-acuity patients discharged by time t, across all nine rules. Although curves remain tightly clustered overall, FCFS underperforms during the first five hours yet surpasses many strategies at later thresholds, mirroring the tail-behavior patterns identified in Section 3.1.

We now present the AUC metrics, formally defined in Appendix D, in Table 3. In addition to the standard AUC computed over a 12-hour window, we introduce three complementary variants: (1) a half-range AUC limited to the first 6 hours, (2) a weighted AUC emphasizing earlier thresholds via a linearly decreasing weight function, and (3) a weighted AUC emphasizing later thresholds via a linearly increasing weight function. These variants enable us to evaluate whether the choice of AUC definition alters performance conclusions.

**Table 3.** Area-Under-Curve (AUC) Metrics for Threshold-Based Discharge Curves. . This table reports four AUC variants—standard (0–12 h), half-range (0–6 h), early-emphasis (linearly decreasing weights), and late-emphasis (linearly increasing weights)—for low- and mid-acuity cohorts under each strategy. Despite different weighting schemes, rankings remain largely consistent; Acuity-Based FCFS appears more favorable under early-emphasis, reflecting its strong initial discharge rates.

| Group | Strategy | AUC (12h) | AUC (6h) | AUC (low-emphasis) | AUC (high emphasis) |
| --- | --- | --- | --- | --- | --- |
| Low | FCFS | 76% | 53% | 59% | 94% |
| Low | APQ | 74% | 49% | 57% | 93% |
| Low | LWP | 74% | 49% | 56% | 93% |
| Low | AAPQ-PFT | 74% | 48% | 56% | 92% |
| Low | AAPQ | 74% | 48% | 56% | 92% |
| Low | Acuity-Based FCFS | 74% | 48% | 56% | 92% |
| Low | PFT | 74% | 47% | 55% | 92% |
| Low | AAPQ-LWP | 73% | 47% | 55% | 92% |
| Low | AAPQ-LWP-PFT | 73% | 47% | 55% | 92% |
| Mid | AAPQ-LWP-PFT | 74% | 49% | 57% | 92% |
| Mid | AAPQ-LWP | 73% | 48% | 56% | 91% |
| Mid | AAPQ | 72% | 45% | 54% | 90% |
| Mid | LWP | 72% | 45% | 53% | 91% |
| Mid | AAPQ-PFT | 70% | 43% | 52% | 89% |
| Mid | PFT | 70% | 42% | 51% | 90% |
| Mid | Acuity-Based FCFS | 70% | 42% | 51% | 89% |
| Mid | APQ | 68% | 39% | 49% | 88% |
| Mid | FCFS | 63% | 29% | 42% | 86% |

Overall, the conclusions remain stable across AUC definitions. One exception is Acuity-Based FCFS, which appears more favorable under the variant emphasizing early thresholds—consistent with Figure 1, where it dominates in the initial portion of the curve.

Next, we sort strategies from best to worst based on each metric. This confirms earlier trends: FCFS ranks highest for low-acuity patients but lowest for mid-acuity patients, whereas AAPQ-LWP-PFT achieves the opposite pattern, performing best in the mid-acuity group but worst in the low-acuity group.

Finally, we quantify each strategy’s responsiveness in Table 4 by reporting the minimum LOS threshold required to serve 50%, 75%, 90%, and 95% of patients. These metrics respectively correspond to the time needed to reach the median, upper-quartile, near-complete, and very-high service-level benchmarks. Together, they provide a granular view of how quickly each strategy meets increasingly stringent performance goals. We find that the choice of threshold meaningfully affects the relative ranking of some strategies but not others. For instance, in the low-acuity cohort, AAPQ-PFT reaches 50% of patients within 185 minutes—midway between FCFS (170 min) and Acuity-Based FCFS (200 min). However, to reach 95%, AAPQ-PFT requires 315 minutes, which is far closer to the worst-performing Acuity-Based FCFS (325 min) than to the best-performing FCFS (260 min). In contrast, strategies like AAPQ-LWP-PFT in the mid-acuity cohort demonstrate consistently strong performance across all thresholds, making them less sensitive to the choice of benchmark.

**Table 4.** Minimum LOS Thresholds to Achieve Service-Level Benchmarks. . This table lists, for each strategy and cohort, the minimum length-of-stay threshold required to discharge 50%, 75%, 90%, and 95% of patients. These responsiveness metrics reveal how quickly each rule meets progressively stringent performance targets, highlighting strategy sensitivity to the chosen benchmark.

| Group | Strategy | 50% | 75% | 90% | 95% |
| --- | --- | --- | --- | --- | --- |
| Low | FCFS | 170 | 200 | 235 | 260 |
| Low | LWP | 190 | 220 | 260 | 290 |
| Low | APQ | 180 | 220 | 265 | 315 |
| Low | AAPQ-LWP | 195 | 230 | 270 | 305 |
| Low | AAPQ-LWP-PFT | 195 | 230 | 270 | 305 |
| Low | AAPQ-PFT | 185 | 230 | 270 | 315 |
| Low | PFT | 190 | 230 | 270 | 310 |
| Low | AAPQ | 190 | 230 | 275 | 310 |
| Low | Acuity-Based FCFS | 200 | 255 | 295 | 325 |
| Mid | AAPQ-LWP-PFT | 175 | 235 | 305 | 375 |
| Mid | LWP | 190 | 250 | 310 | 365 |
| Mid | AAPQ-LWP | 180 | 240 | 325 | 400 |
| Mid | PFT | 205 | 265 | 330 | 380 |
| Mid | AAPQ | 195 | 255 | 335 | 395 |
| Mid | AAPQ-PFT | 200 | 270 | 350 | 415 |
| Mid | APQ | 220 | 280 | 360 | 410 |
| Mid | Acuity-Based FCFS | 210 | 275 | 360 | 420 |
| Mid | FCFS | 260 | 320 | 370 | 395 |

### 3.3 Stakeholder-informed utility analysis

We evaluate two utility functions—an elliptical form, *U*_1_, and a linear form, *U*_2_(definitions in Appendix B)—by first plotting their values against the LOS threshold *t*. Figures 3 and 4 display the curves *U*_1_( *T*_LOS―Low_(*t*), *T*_LOS―Mid_(*t*)) and *U*_2_(*T*_LOS―Low_(*t*), *T*_LOS―Mid_(*t*)), respectively, allowing us to observe how strategy rankings evolve as *t* increases. Because both curves preserve the same ordering up to approximately five hours, the choice of threshold has minimal impact on the relative performance of the rules—whereas frequent crossings would mandate consideration of multiple thresholds to capture divergent conclusions.

**Figure 3.** Elliptical Utility U_1_ Across Thresholds. This plot shows the elliptical utility U_1_(T_LOS―Low_(t), T_LOS―Mid_(t)) for each strategy as a function of the discharge-time threshold t. Consistent vertical ordering up to approximately five hours indicates that strategy rankings under U_1_ are robust to the choice of t within this range.

**Figure 4.** Linear Utility U_2_ Across Thresholds. This figure depicts the linear utility U_2_(T_LOS―Low_(t),T_LOS―Mid_(t)) plotted against t. The near–parallel curves for most rules confirm that, under U_2_, conclusions about relative strategy performance remain largely unaffected by threshold selection.

We then fix a threshold *t*^∗^ and generate scatter plots of (*T*_LOS―Low_(*t*^∗^), *T*_LOS―Mid_(*t*^∗^)) overlaid with utility contours *U*( · ) = *c*, using color to encode utility level. In Figures 5 and 6, which employ the elliptical utility *U*_1_, we set *t*^∗^ = 5h and 7h. At five hours, FCFS achieves the highest low-acuity discharge rate but ranks poorly overall because its mid-acuity rate trails the next-worst strategy (APQ) by roughly 12 percentage points, resulting in low utility. The other strategies differ only marginally on low-acuity performance, with mid-acuity rates spanning about nine points between APQ and AAPQ-LWP-PFT. At seven hours, FCFS’s utility improves dramatically—rising from worst to second best—illustrating how deeper tail performance can overturn short-threshold conclusions. Figures 7 and 8 repeat this analysis with the linear utility *U*_2_, confirming that FCFS’s strong long-threshold performance persists despite its weaker results at shorter thresholds.

**Figure 5.** Elliptical Utility Contours at t^∗^ = 5 hours. Scatter plot of (T_LOS―Low_(5h),T_LOS―Mid_(5h)) for each strategy, overlaid with elliptical-utility contours U_1_ = c. At this threshold, FCFS maximizes low-acuity discharges but scores low overall due to substantially poorer mid-acuity performance, illustrating the trade-off captured by U_1_.

**Figure 6.** Elliptical Utility Contours at t^∗^ = 7 hours. Scatter plot of (T_LOS―Low_(7h),T_LOS―Mid_(7h)) with U_1_ contours. At seven hours, FCFS moves from the worst to the second-best position, demonstrating how right-tail evaluation alters strategy rankings under the elliptical utility.

**Figure 7.** Linear Utility Contours at t^∗^ = 5 hours. Scatter of (T_LOS―Low_(5h),T_LOS―Mid_(5h)) with linear-utility contours U_2_ = c, the linear utility penalizes strategies with imbalanced performance, reaffirming FCFS’s disadvantage at shorter thresholds.

**Figure 8.** Linear Utility Contours at t^∗^ = 7 hours. Scatter of (T_LOS―Low_(7h),T_LOS―Mid_(7h)) with U_2_ contours. The shift in FCFS’s relative position—from low to high utility—mirrors the pattern seen under the elliptical utility, underscoring consistency across utility forms.

Finally, Figure 9 compares *U*_1_ and *U*_2_at *t*^∗^ = 3h by plotting each strategy’s pair of utility values. The near-monotonic alignment along the regression line indicates that the choice of utility form and hyperparameters exerts minimal influence on strategy ranking; a more scattered pattern would have revealed sensitivity to the utility specification.

**Figure 9.** Comparison of Elliptical and Linear Utilities at t^∗^ = 7 hours. Bivariate plot of each strategy’s utility values (U_1_,U_2_) at t^∗^ = 3h, with a fitted regression line. The near-monotonic alignment indicates minimal sensitivity of strategy rankings to the choice of utility function or its hyperparameters.

### 3.4 Strategy Strengths and Weaknesses

Table 5 synthesizes each strategy’s principal strengths and weaknesses, with supporting tables or figures indicated in parentheses. We reiterate that the aim of this analysis is to demonstrate our evaluation framework and to illustrate how it can uncover strategy-specific trade-offs—not to endorse any particular rule. These findings should not be used as implementation guidance, since they are based on a single simulation scenario and on illustrative utility parameters rather than stakeholder-calibrated values.

**Table 5.** Strategy Strengths, Weaknesses, and Deployment Recommendations. This table summarizes each prioritization rule’s principal performance advantages and drawbacks—citing relevant analysis panels (T2–T4 refer to Tables 2–4; F1–F9 refer to Figures 1–9)—and provides a high-level recommendation for use. Strengths highlight metrics or utility benchmarks where a strategy excels, while weaknesses identify its shortcomings. Recommendations classify each rule as “Strong contender,” “Acceptable,” “Conditional,” or “Avoid,” based on the balance of benefits and implementation trade-offs under the illustrative simulation scenario.

| Strategy | Strengths | Weaknesses | Recommendation |
| --- | --- | --- | --- |
| FCFS | Fastest low-acuity mean, median, and upper tail (T2)<br>Best mid-acuity 99th percentile (T2)<br>Outstanding low-acuity discharge rates and AUC (F1; T3) | Slow early tail and central metrics for mid-acuity (T2)<br>Poor mid-acuity discharge rates and AUC (F2; T3)<br>Very low stakeholder utility until late thresholds (F3–F9) | **Avoid.** Prioritizes minor cases at the expense of higher-acuity patients, yielding substandard performance for mid-acuity despite strong low-acuity results. |
| Acuity-Based FCFS | Lowest absolute maximum LOS (T2)<br>Highest low-acuity discharge rate in first 2 h (F1) | Worst low-acuity median and upper percentiles (T2)<br>Worst mid-acuity 95th percentile (T2)<br>Mediocre on most LOS metrics (T2) | **Eliminate.** Although intuitively fair, it underperforms for both acuity groups across nearly all metrics. |
|  |  | • Low utility across thresholds (F3–F9) |  |
| APQ | • Strong low-acuity central tendencies and 75<sup>th</sup> percentile (T2)<br>• Solid overall AUC (T3) | • Worst low-acuity 99th percentile and poor 95th percentile (T2)<br>• Weak mid-acuity LOS and discharge times (T2; T4)<br>• Lower utility relative to alternatives (F5–F9) | **Avoid.** Limited benefits for low-acuity do not justify its significant mid-acuity and tail drawbacks. |
| PFT | • Excellent mid-acuity tail performance (95th, 99th, STD) (T2)<br>• Fast time to 95% mid-acuity discharge (T4)<br>• Top utility at 7 h thresholds (F6; F8) | • Below-average low-acuity mean (T2)<br>• Subpar overall AUC (T3) | **Acceptable.** Mid-acuity strengths and strong late-threshold utility offset modest low-acuity and AUC weaknesses. |
| LWP | • Above-average LOS statistics for both cohorts (T2)<br>• Good mid-acuity AUC and discharge times (T3; T4)<br>• Best 5 h utility (F5; F7) | • Average low-acuity AUC and median discharge (T3; T4)<br>• Mid-acuity 99th percentile only moderate (T2) | **Strong contender.** Consistently solid performance with few notable weaknesses. |
| AAPQ | • Balanced central tendencies and moderate tails for both cohorts (T2)<br>• Good low-acuity AUC (T3)<br>• Utility at or above average (F5–F9) | • Below-average mid-acuity AUC (T3)<br>• Slower low-acuity threshold attainment (T4) | **Acceptable.** Not the top performer but offers a good balance between low- and mid-acuity outcomes. |
| AAPQ-PFT | • Above-average low-acuity 50% discharge time (T4) | • Below-average low-acuity 95th/99th percentiles (T2)<br>• Weak mid-acuity 90th/95th percentiles (T2)<br>• Low utility at common thresholds (F5–F8) | **Avoid.** Combines two moderate strategies but underperforms each individually. |
| AAPQ-LWP | • Top mid-acuity central tendencies and 75th percentile (T2)<br>• High mid-acuity AUC (T3)<br>• Excellent mid-acuity 50%/75% discharge rates (T4) | • Worst low-acuity mean and 99th percentile (T2)<br>• Low low-acuity AUC (T3)<br>• Worst mid-acuity 99th percentile (T2) | **Conditional.** Best for improving mid-acuity LOS but imposes substantial low-acuity and tail trade-offs. Stakeholder priorities must guide its use. |
|  | • Strong utility at 3 h and 5 h (F5; F7; F9) |  |  |
| AAPQ-LWP-PFT | • Best mid-acuity central, 75th/90th percentiles, AUC, and discharge times (T2; T3; T4)<br>• Among highest utility values across thresholds (F5–F9) | • Worst low-acuity mean and AUC (T2; T3)<br>• Slow low-acuity 50% discharge (T4) | **Acceptable.** Excels for mid-acuity and overall utility; low-acuity delays may be acceptable if stakeholders focus on higher-acuity patient flow. |

## 4 Discussion

### 4.1 Synthesis of principal findings

The results presented in Section 3 collectively demonstrate that the choice of evaluation lens—summary statistics, threshold-attainment curves, or stakeholder-weighted utilities—materially shapes how ED prioritization strategies are ranked. Even within a single 30-bed simulated ED, a rule that looks benign under a LOS comparison can exhibit sizeable right-tail liabilities, while another rule that excels on extreme percentiles may fall behind when stakeholder utilities emphasize shorter thresholds. These contrasts underscore the central argument of this study: no single metric is sufficient to capture the multidimensional consequences of crowd management policies, and conclusions should never be drawn from a solitary point estimate.

### 4.2 Why cohort stratification is indispensable

Averaging performance across acuity bands smooths precisely the variation clinicians and administrators most need to see. When low- and mid-acuity LOS are pooled, the observed spread between strategies narrows to just 7 minutes, suggesting no meaningful distinction. However, when evaluated separately, the spread expands to 22 minutes for low-acuity patients and 74 minutes for mid-acuity patients—highlighting substantial and actionable differences in strategy performance.

By diluting the average LOS in this fashion, an analyst might incorrectly infer that any of the nine rules is “good enough,” missing the variation for each group of patients. Operationally, such masking can divert resources toward the wrong bottleneck or delay recognition of inequitable wait times among patient groups.

### 4.3 Strategic trade-offs across patient groups

Ideal policies would compress the entire KPI distributions for every cohort, yet the data reveal persistent trade-offs. FCFS and AAPQ-LWP-PFT epitomize this tension: the former minimizes low-acuity mean and upper-tail LOS, whereas the latter delivers the best mid-acuity central and tail performance at the expense of longer waits for low-acuity cases.

These findings reinforce the need for hospitals to state explicit priorities—whether reducing crowding for mid-acuity patients, accelerating flow for discharged fast-track patients, or balancing both objectives through mixed strategies or dynamic rules.

### 4.4 Statistical significance versus clinical relevance

Table 2 shows that the worst low-acuity mean LOS (192 min) is twenty-two minutes longer than the best (170 min). While statistically significant, administrators must decide whether a twenty-minute average difference is operationally meaningful when the clinically acceptable window for discharge may span several hours. Hence, statistical tests alone cannot substitute for clinical judgment; practical significance must be interpreted in context.

### 4.5 Actionability of threshold-attainment metrics

Decision makers consistently ask, “How quickly can we reach a given service target?” Table 4 answers this in language executives and frontline clinicians immediately understand. For instance, APQ discharges half of low-acuity patients twenty minutes sooner than Acuity-Based FCFS, and it reaches the 90 percent benchmark thirty minutes earlier. Such statements translate directly into staffing discussions, fast-track lane designs, and patient-communication scripts.

### 4.6 Insights from strengths–weaknesses mapping

The qualitative synthesis in Table 5 distills dozens of numerical comparisons into a concise narrative of strategic profiles. Strikingly, all identified strengths and weaknesses stem from a single KPI—LOS—in one virtual ED. If this level of heterogeneity arises from one performance dimension, the variation across additional KPIs such as left-without-being-seen rates, bed-blocking time, or door-to-doctor-time is likely far greater. The implication is sobering: past studies that benchmarked policies on a single mean LOS or four-hour target may have drawn overly general conclusions, exacerbating translation gaps when rules are transplanted to new hospitals.

### 4.7 Central role of sensitivity analysis

Throughout Section 3, sensitivity analysis served as a safeguard against misleading conclusions. For instance, Figures 5 and 6 displayed the Elliptical utility function evaluated at five- and seven-hour thresholds, respectively. Despite the modest two-hour difference, the resulting strategy rankings shifted noticeably. FCFS, which appeared to perform worst under the five-hour threshold, ranked among the best when evaluated at seven hours. Relying on a single threshold would have led to a threshold-dependent— and potentially distorted—interpretation of FCFS performance. In contrast, other metrics, such as AUC-to-time-to-threshold, demonstrated robustness to parameter choices, producing consistent results across variations. Nonetheless, systematically conducting sensitivity analyses remains helpful to guard against occasional but consequential parameter-driven artifacts. Recognizing these dependencies discourages overconfidence in model generalizability and helps ensure that operational insights remain valid when applied to alternate settings.

### 4.8 Recommended Steps for Strategy Evaluation

To ensure that patient-prioritization studies are both rigorous and transparent, we offer the following guidelines for researchers. These principles will help one apply each evaluation technique consistently, highlight strategy trade-offs, and guard against overconfident claims of “best” performance.

Technique 1 should be employed across all KPIs and cohorts by reporting sample size, mean, median, minimum, maximum, and key upper-tail percentiles (75th, 90th, 95th, 99th) for each clinically relevant subgroup (e.g., acuity level, arrival window, resource requirement, boarding status). Technique 2 (threshold-attainment) ought to accompany every KPI, with concise tabular summaries of the time required to reach a stakeholder-defined attainment level (such as 90 percent) for each cohort, thus providing both clinical relevance and ease of interpretation. Technique 3 (stakeholder-informed utility) should then translate the most critical KPI(s) into a single utility score that incorporates stakeholder preferences— thereby facilitating equitable, real-world comparisons. Throughout, results should be presented as context-dependent trade-offs rather than absolute winners, with each strategy’s strengths and weaknesses clearly articulated across techniques and cohorts; universal superiority should be asserted only when an exhaustive range of KPIs, cohorts, evaluation methods, and operational scenarios has been rigorously tested and validated.

We conclude by reflecting on how often each evaluation technique was cited in our analysis summary in Section 3.4 (Table 5). We counted the number of times Techniques 1–3 were referenced when comparing each strategy’s strengths and limitations. Technique 1 (detailed summary statistics) accounted for only 26% of all citations—yet most researchers report only a less-detailed subset of these metrics, implying much information about upper-tail behavior and cohort-specific performance is routinely omitted. Technique 2 (threshold attainment) and Technique 3 (stakeholder-informed utility) comprised roughly 29% and 45% of references, respectively. While citation frequency does not directly imply importance, it offers a useful proxy for how readily each approach reveals key trade-offs—underscoring the need to apply all three in concert rather than relying on any single method.

### 4.9 Limitations and directions for future research

We reiterate that the purpose of this study is to introduce and demonstrate a suite of evaluation techniques— not to endorse any particular prioritization strategy. Several limitations caution against over-interpreting the illustrative results. The simulation was calibrated to a single ED with 30 beds and moderate patient volume. As such, different acuity distributions, arrival patterns, boarding durations, or staffing constraints in other settings may yield materially different rankings. Moreover, our analysis focused exclusively on LOS, omitting other important outcomes such as door-to-doctor time, patient safety, and staff workload. The utility parameters used in this study were intended for illustrative purposes only; they were not elicited from stakeholders or validated through multicenter analysis.

This paper introduces three evaluation techniques designed to promote more rigorous and transparent comparisons of patient prioritization strategies. While these techniques provide a structured foundation, they are not comprehensive. Future work should aim to develop additional methods that illuminate distinct strengths and weaknesses of each strategy. Although quantitative tools such as AUC can be useful, their benefits are limited if they lack interpretability or practical relevance. When mathematical measures are employed, visualizations should be designed to clearly convey real-world implications—as exemplified by the utility function plots in Figures 5 through 8.

Our evaluation framework was applied to a single ED configuration, making it well suited for guiding local implementation decisions. However, many researchers aim to develop strategies that generalize across multiple ED settings. The most direct approach to support such generalization is to apply the evaluation techniques across a diverse set of simulated ED environments—ideally spanning 4 to 8 configurations that vary by bed count, staffing model, and arrival volume. While this approach provides valuable insights, it is resource-intensive: even a single-configuration analysis across multiple KPIs can span dozens of pages. A promising direction for future research is to identify principled methods for aggregating results across configurations. One such approach, based on covariance ovals, is introduced in Appendix B.5. Care must be taken to preserve between-scenario variability, as aggregation can obscure meaningful differences. For example, in Section 3.1, combining low- and mid-acuity cohorts reduced the mean LOS difference across strategies from 22 and 74 minutes, respectively, to only 7 minutes—thereby masking clinically relevant distinctions.

Although the proposed techniques allow rigorous numerical comparisons of patient-prioritization strategies, implementation feasibility must be assessed *before* any analysis begins. A reinforcement-learning (RL) policy (e.g., see Lee and Lee [27]), for example, may offer superior theoretical performance, yet its practical requirements can be prohibitive. Many RL formulations presume real-time knowledge of each patient’s location, resource needs, and even uncertain variables such as future arrivals and service times. Collecting this information in simulation is straightforward; acquiring it in a working ED would demand either extensive manual data entry—adding workload for clinical staff—or continuous computer-vision monitoring, which entails new infrastructure, software, and privacy concerns. A strategy that reduces waiting times on paper but imposes substantial operational or ethical costs provides little real benefit. Researchers should therefore verify that any recommended policy can be deployed with existing data streams and minimal additional burden on personnel; otherwise, theoretical optimality will remain purely academic.

Finally, we briefly reflect on the performance of our novel strategies—PFT, LWP, AAPQ, and their combinations—within the illustrative analysis. While we emphasize that these results are not intended as definitive endorsements, it is noteworthy that our proposed strategies consistently outperformed traditional benchmarks such as Acuity-Based FCFS and APQ across multiple evaluation criteria. As such, these strategies represent a secondary contribution to this work: they appear promising and merit further investigation. In addition to their favorable quantitative performance, they were deliberately designed for ease of implementation and interpretability, making them attractive candidates for real-world adoption.

## 5 Conclusion

This study advances the science of ED operations by formalizing and empirically demonstrating a tripartite evaluation framework—tail-sensitive summary statistics, threshold-attainment profiles, and stakeholder-informed utility analysis—for transparent comparison of patient-prioritization strategies. Applying the framework to nine rules within a common discrete-event simulation revealed that strategic rankings are highly sensitive to the chosen evaluative lens and to cohort aggregation: analyses confined to overall means understated clinically meaningful differences that emerged once low- and mid-acuity patients were considered separately, and strategies that appeared dominant under central-tendency metrics were often eclipsed when extreme percentiles or utility-weighted outcomes were examined. These findings confirm that no single KPI, time target, or composite score adequately captures the multidimensional consequences of queue-management decisions, and they underscore the practical necessity of reporting a minimum set of distributional, threshold-based, and preference-aligned measures.

By framing results as trade-offs rather than pronouncing universal “winners,” the proposed framework equips hospital leaders to align prioritization rules with explicit local objectives—whether accelerating flow for mid-acuity patients, protecting low-acuity throughput, or balancing both via mixed or adaptive policies. Because all three techniques rely on familiar statistical constructs and easily visualized curves, the methodology remains accessible to multidisciplinary stakeholders while satisfying the rigor demanded by academic investigators and quality-improvement teams. Moreover, the framework is KPI-agnostic and extensible to additional outcomes such as door-to-doctor time, boarding duration, patient safety indicators, or staff workload, inviting comprehensive performance audits without presupposing any specific metric hierarchy.

Several limitations temper the generalizability of the illustrative results. The simulation reflected a single, moderately busy, 30-bed ED; different capacity profiles, arrival patterns, or boarding pressures may yield alternative strategic orderings. Only length of stay was modeled, utility parameters were illustrative rather than elicited, and no external validation across multiple health systems was undertaken. Future research should therefore replicate the framework across diverse ED configurations, incorporate a broader KPI portfolio, elicit context-specific utilities, and test whether adaptive or hybrid strategies can dominate static rules when assessed under the full triad of metrics. Methodological work is also needed to synthesize results across multiple scenarios without obscuring cross-site heterogeneity—an aggregation challenge analogous to cohort masking within a single ED.

Notwithstanding these caveats, the present contribution provides a replicable template that reconciles statistical robustness with managerial interpretability, furnishing researchers and practitioners with a common language for evidence-based policy design. Widespread adoption of this evaluation standard promises to accelerate meta-analysis, clarify when and where novel prioritization algorithms add value, and ultimately promote safer, timelier, and more equitable emergency care.

## Data Availability

We made the data and code publicly available on the following GitHub repository: https://github.com/adamdeho/ER-Evaluation-Techniques.git.

---

S1 Appendix A. Patient Prioritization Strategies

S1 Appendix B. Utility Functions

S1 Appendix C. Discrete Event Simulation Details

S1 Appendix D. Variants of the Area Under the Curve Metric

